# 3D-DXA-based Finite Element Models Improve Hip Fracture Prediction versus aBMD in the Japanese Population-Based Osteoporosis (JPOS) Cohort Study

**DOI:** 10.64898/2026.11.16.26368177

**Authors:** Elham Alizadeh, Junko Tamaki, Abdollah Moslemi, Takahiro Tachiki, Yuho Sato, Masami Hamada, Kouji Tsuda, Asako Kudo, Kuniyasu Kamiya, Nami Imai, Akemi Nitta, Kumiko Ohara, Naoyuki Takashima, Jun Kitagawa, Katsuyasu Kouda, Akemi Morita, Kazuhiro Uenishi, Etsuko Kajita, Sadanobu Kagamimori, Ludovic Humbert, Jérôme Noailly, Masayuki Iki

## Abstract

**Summary:** Hip fractures are among the most serious consequences of osteoporosis, yet many individuals who sustain hip fractures do not meet the diagnostic threshold based on areal bone mineral density (aBMD). Finite element (FE)-based mechanical descriptors may improve hip fracture prediction beyond aBMD.

**Purpose:** This study investigated whether FE-based mechanical descriptors derived from dual-energy X-ray absorptiometry (DXA) improve hip fracture discrimination beyond aBMD in the Japanese Population-Based Osteoporosis (JPOS) cohort.

**Methods:** The study included 1,636 women, of whom 85 sustained hip fractures during 20 years of follow-up. Subject-specific three-dimensional femoral models were reconstructed from DXA images using statistical shape and appearance modelling, and nonlinear FE analyses were performed to derive mechanical descriptors under simulated sideways-fall loading. Predictive performance was evaluated using discrimination, model fit, reclassification, and hazard ratio analyses.

**Results:** Over two-thirds of women who experienced hip fracture were not classified as osteoporotic based on aBMD. Several FE-based descriptors significantly improved fracture prediction relative to total hip and femoral neck aBMD across the evaluation frameworks. The best-performing descriptor, combining femoral strength with residual energy (*F*_*U*_ + *W*_*R*_), achieved a significantly higher AUROC of 0.797 compared with 0.763 (p = 0.026) and 0.757 (p = 0.040) for total hip and femoral neck aBMD, respectively, and the highest hazard ratio (3.577 per SD). The same descriptors consistently performed best across all evaluation methods.

**Conclusion:** These findings demonstrate that FE-based descriptors provide meaningful information beyond aBMD and may improve identification of women at elevated hip fracture risk.

## Introduction

Osteoporosis is a chronic condition characterized by low bone mass and reduced bone microstructural integrity, increasing skeletal fragility and fracture risk with aging [1], with hip fractures imposing the greatest socioeconomic burden among fragility fractures [2]. Fracture prevention requires identifying individuals at high risk, typically through areal bone mineral density (aBMD) measured by dual-energy X-ray absorptiometry (DXA), with T-scores indicating deviations from healthy young adults. However, more than 50% of incident hip fractures occur in women without osteoporosis (aBMD T-score > −2.5) [3]. Consequently, reliance on aBMD alone for fracture risk prediction does not adequately identify individuals at increased risk, as the mechanical competence of the proximal femur depends not only on bone mineral density but also on a broader range of organ- and subject-specific characteristics.

Hip fracture risk is associated with reduced femoral strength, influenced by aging, aBMD, bone geometry, cortical and trabecular microarchitecture and anisotropy, and the magnitude and direction of mechanical loading. Accordingly, image-based, subject-specific finite element models (FEM) have been used to estimate femoral strength and assess fracture resistance [4–6]. While initial studies reported no added predictive value of femoral strength over aBMD [7, 8], more recent studies have demonstrated the superiority of FEM-derived measures [5, 9]. However, most of these approaches rely on quantitative computed tomography (QCT) [4, 5], which involves higher radiation exposure and costs, limiting its routine use in osteoporosis screening and longitudinal fracture risk assessment [10].

DXA is routinely used for osteoporosis screening due to its low radiation exposure and cost. Several studies have applied DXA-based 2D FEM to predict femoral strength and fracture risk in clinical cohorts [11–13]. Although these models capture aspects of femoral mechanical behavior [11], their performance in osteoporosis-related fracture classification remains limited [13]. A key limitation is the lack of volumetric information, preventing accurate representation of out-of-plane variations in femoral geometry and bone mineral density.

The field subsequently explored DXA-based 2D-to-3D reconstruction techniques for developing 3D FEM [14, 15]. Grassi et al. used statistical shape and appearance modeling (SSAM) to reconstruct subject-specific 3D femoral anatomy from DXA images [16] and perform FE simulations [6, 17]. Using this approach, studies in the MrOS male cohort [6] and OSTPRE female cohort [17] demonstrated that FEM-derived bone strength, normalized to body weight and evaluated across 10 side-fall loading configurations, significantly improved hip fracture discrimination compared with TH aBMD and FN aBMD, respectively, adjusted for BMI. Despite these improvements, clinical implementation remains limited by computational costs [5], the need for greater automation [18], and the absence of regulatory approval [6]. Furthermore, most approaches rely primarily on bone strength, typically defined as the maximum reaction force, or for Grassi’s model [6], evaluated under an external load model for side-fall. Incorporating additional FE-based descriptors of deformability and mechanical energy absorption could provide complementary information on bone fragility and improve fracture discrimination [19].

The 3D-Shaper® software (v2.14, 3D-Shaper Medical, Barcelona, Spain) uses a computationally efficient SSAM-based approach to reconstruct subject-specific 3D femoral geometry and volumetric bone mineral density distributions from DXA images [14]. The software is commercially available and has received regulatory clearance in several territories, including the European Union, United States, Japan, and Brazil. A 3D-DXA-based FE pipeline incorporating 3D-Shaper was developed to estimate femoral mechanical response under subject-specific loading [20], with femoral strength estimates evaluated against QCT-based models [20, 21]. Retrospective case–control studies in two Spanish cohorts demonstrated that FE-based mechanical descriptors provide complementary information to BMD for hip fracture discrimination [19, 22]. More recently, a prospective study in the Geneva Retirees Cohort [23] reported comparable associations of femoral strength and TH aBMD with major osteoporotic fractures (MOF), while a strength threshold of 2600 N improved fracture risk classification compared with TH aBMD-based thresholds. Notably, 27% of women with MOF who were not classified as osteoporotic by aBMD exhibited reduced femoral strength, supporting its complementary value in fracture risk assessment.

To date, the Geneva Retirees Cohort is the only prospective study [23] evaluating the aforementioned 3D-DXA-based FEA [20], highlighting the need for further validation in larger and more diverse populations with more hip fracture events. Accordingly, the present study applies our previously developed nonlinear 3D-DXA-based FEM [20] to the prospective, population-based Japanese Population-Based Osteoporosis (JPOS) cohort to evaluate the predictive value of FE-based mechanical descriptors for hip fracture risk. These descriptors, introduced in our previous work [19], integrate femoral geometry, bone density distribution, mechanical properties, and tissue-dependent structural characteristics, including measures of accumulated plastic strain up to femoral strength. To our knowledge, this is the first study to investigate whether FE-based mechanical descriptors derived from standard hip DXA scans outperform aBMD for hip fracture prediction in a prospective, population-based cohort of women.

## Materials and Methods

### Study cohort and DXA data

The study population was derived from the Japanese Population-Based Osteoporosis (JPOS) study, as previously described [24]. Briefly, women aged 15–79 years were randomly sampled in 5-year age strata from seven regions across Japan, five of which were included in the JPOS Cohort Study. The cohort comprised 1,636 women, including 85 who subsequently sustained a hip fracture. Participants completed the baseline assessment and were followed for up to 20 years through up to five surveys.

### 2D-to-3D reconstruction

3D-Shaper software was used to reconstruct subject-specific 3D models of the proximal femur from standard hip DXA scans [14]. The method uses an SSAM derived from a QCT database, with femoral shape and volumetric density variability represented by principal component analysis. Through iterative 3D–2D registration, shape, density, and pose parameters are optimized to match the model-derived projection to the DXA image, reconstructing both cortical and trabecular compartments.

The resulting measurements include integral vBMD (Int.vBMD, mg/cm^3^): mean vBMD across cortical and trabecular compartments in the total hip (TH) region; trabecular vBMD (Trab.vBMD, mg/cm^3^): mean vBMD within the TH trabecular compartment; and cortical surface BMD (Cort.sBMD, mg/cm^2^): calculated as cortical vBMD multiplied by cortical thickness and averaged over the TH region. These measurements have been validated against QCT [14, 21, 25].

### Finite element modeling

The previously developed 3D-DXA-based FE framework [19, 20] was used to simulate femoral mechanical behavior under sideways fall loading. Subject-specific FE models were generated by morphing a generic mesh of linear eight-node hexahedral elements to the 3D-Shaper-reconstructed femoral geometry using thin-plate spline (TPS) morphing, preserving cortical and trabecular compartments. The reconstructed bone density distribution was mapped onto the mesh by linear interpolation. Compartment-specific material properties were assigned using established density– elasticity relationships [19], converting 3D-DXA-derived vBMD to ash density and then to apparent density to calculate Young’s modulus. A constant Poisson’s ratio of 0.3 was assumed. Nonlinear behavior was modeled using an elastic–perfectly plastic constitutive model with density-dependent, tension- and compression-specific yield properties for cortical and trabecular bone [19].

Boundary conditions reproduced an experimental lateral fall configuration [11], and widely adopted in FE-based femoral strength analyses [6], were applied in the FE simulations. The distal shaft was fully constrained, while the greater trochanter was constrained only along the fall direction. Displacement was applied to the femoral head through rigid-body constraints until the reaction force decreased with increasing displacement, indicating a surrogate marker of mechanical failure. All models were simulated with 10° adduction and 15° internal rotation to represent a typical sideways fall in the available experimental setups [26]. The FE pipeline is illustrated in Fig. 1.

**Fig. 1.**
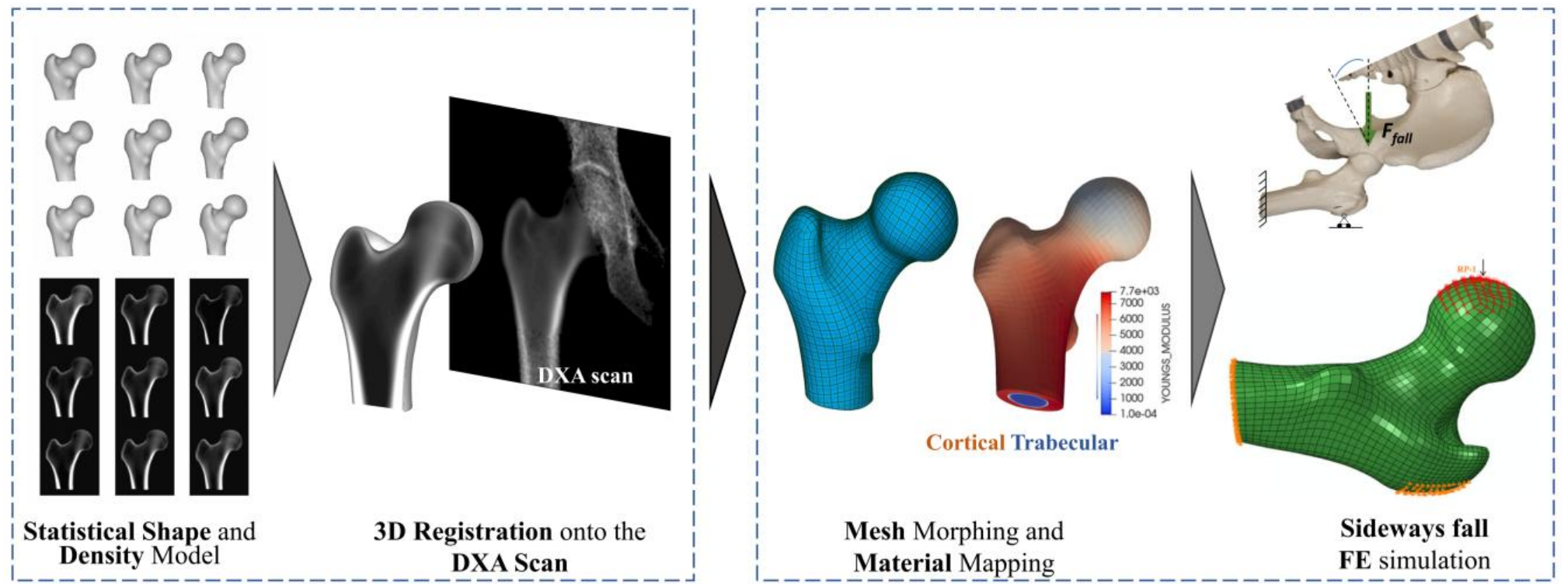
Schematic representation of the methodological framework applied in the present study for fracture prediction.

### Mechanical descriptors

The detailed methodology, mechanical definitions, and clinical relevance of the FE-based descriptors have been described in our previous study [19]. Mechanical descriptors were extracted from the nonlinear FE force–displacement response to characterize femoral strength, deformability, and energy absorption (Supplementary Fig. A1). Primary nonlinear descriptors included strength (*F*_*U*_) (maximum reaction force), displacement at failure (*D*_*U*_), and nonlinear energy to failure (*W*_*U*_) [19]. An optimized piecewise-linear analysis [19] was applied to derive elastic descriptors (linear elastic force (*F*_*LE*_), linear elastic displacement (*D*_*LE*_), and linear elastic energy (*W*_*LE*_)) and post-yield nonlinear deformation (*D*_1_). A loading–unloading analysis was additionally performed on the nonlinear FE models to quantify the mechanical energy dissipated through accumulated plastic deformation, from which three key descriptors were derived: residual displacement (*D*_*R*_), dissipated energy (*W*_*D*_), and residual (elastic) energy (*W*_*R*_).

To assess the complementary contributions of deformation and energy absorption to fracture discrimination, strength (*F*_*U*_) and linear elastic force (*F*_*LE*_) were separately combined with deformation- and energy-based descriptors. All individual and combined descriptors (Supplementary Table B1) were evaluated for hip fracture discrimination in the full cohort.

### Statistical analysis

Detailed statistical methods are provided in Supplementary C and are briefly summarized here. The predictive performance of the 3D-DXA-based FE mechanical descriptors (Supplementary Table B1) was evaluated through group comparisons, fracture discrimination, and survival analyses (Supplementary Fig. C1). Differences between fracture and control subjects were assessed using unpaired t-tests or Mann–Whitney U tests, according to data distribution. Fracture discrimination was evaluated using logistic regression and the area under the receiver operating characteristic curve (AUROC). Multivariable models combining mechanical descriptors were assessed using leave-one-out cross-validation, and AUROC differences were compared using the DeLong test.

Cox proportional hazards models were used to incorporate time-to-fracture information and censoring. Model discrimination was assessed using Harrell’s C-index [27], with paired differences (ΔC) evaluated against reference predictors using two-sided nonparametric bootstrap resampling. Model fit was compared using Akaike’s Information Criterion (AIC) and differences in AIC (ΔAIC) [28]. The additional predictive value of FE-based descriptors beyond TH or FN aBMD was assessed using categorical net reclassification improvement (NRI), integrated discrimination improvement (IDI), and continuous NRI (cNRI) [29, 30]. Risk categories were established using NHANES III reference data [31], and reclassification analyses were performed over the full available follow-up period.

Associations with hip fracture risk were quantified using hazard ratios (HRs) from ridge-penalized Cox models [32]. Predictors were standardized, and HRs were reported per one standard deviation (SD) change. The ridge penalty parameter (λ) was optimized using 10-fold cross-validation, with the largest optimal λ applied across all models. Nonparametric bootstrap resampling was used to estimate 95% confidence intervals (CIs) and p-values for reclassification and HR analyses. All analyses were performed using MATLAB R2021a (MathWorks, Natick, MA, USA).

## Results

In this study, 67.06% (57 subjects) of women with hip fracture were classified as non-osteoporotic based on TH aBMD, whereas only 29.41% (25 subjects) were classified as osteoporotic using FN aBMD.

Baseline characteristics and mechanical descriptors of the study cohort, stratified by hip fracture status, are presented in Table 1. Participants with hip fractures were significantly older and shorter than controls, with no significant differences in weight or BMI. TH and FN aBMD, all 3D-DXA-derived parameters, and FE-based descriptors differed significantly between groups (p < 0.001). Controls exhibited higher force- and energy-related descriptors (*F*_*U*_, *W*_*U*_, *F*_*LE*_, *W*_*LE*_, *W*_*D*_, and *W*_*R*_) and lower deformation-related descriptors (*D*_*U*_, *D*_*LE*_, *D*_1_, and *D*_*R*_), indicating greater load-bearing capacity and energy absorption with reduced deformation.

**Table 1.** Demographic and clinical characteristics, DXA, 3D-DXA-based parameters, and FE-based descriptors by hip fracture status (mean ± SD).

| Measurement |  |  | Parameters | Control | Hip fracture | *P* <sup>\*</sup> |
|---|---|---|---|---|---|---|
| **Demographic and Clinical** |  |  | Number (%) | 1551 (94.8%) | 85 (5.2%) | - |
|  |  |  | Age (Years) | 58.14±10.66 | 69.10±7.56 | 0.001 |
|  |  |  | Height (*cm*) | 150.93±5.91 | 147.93±5.77 | 0.001 |
|  |  |  | Weight (*kg*) | 54.35±8.32 | 53.20±9.09 | 0.215 |
|  |  |  | BMI ($\frac{kg}{m^2}$) | 23.86±3.38 | 24.25±3.53 | 0.296 |
| **DXA** |  |  | TH aBMD ($\frac{g}{cm^2}$) | 0.804±0.13 | 0.685±0.09 | <0.001 |
|  |  |  | FN aBMD ($\frac{g}{cm^2}$) | 0.708±0.12 | 0.603±0.08 | <0.001 |
| **3D-DXA-based Parameters (TH)** |  |  | Int.vBMD ($\frac{mg}{cm^3}$) | 288.18±61.06 | 230.97±41.46 | <0.001 |
|  |  |  | Trab.vBMD ($\frac{mg}{cm^3}$) | 161.75±43.04 | 120.12±27.54 | <0.001 |
|  |  |  | Cort.sBMD ($\frac{mg}{cm^2}$) | 139.15±24.33 | 118.49±17.75 | <0.001 |
| **FE-Based Descriptors (TH)** | **Nonlinear FE Analysis** |  | *F*<sub>*U*</sub> (N) | 2727.92±671.43 | 2189.92±430.10 | <0.001 |
|  |  |  | *D*<sub>*U*</sub> (mm) | 2.82±0.31 | 3.06±0.30 | <0.001 |
|  |  |  | *W*<sub>*U*</sub> (N.mm) | 5559.90±1249.29 | 4888.89±1067.79 | <0.001 |
|  | **Piecewise Linear Analysis** | *Elastic* | *F*<sub>*LE*</sub> (N) | 2203.39±554.05 | 1747.35±348.42 | <0.001 |
|  |  |  | *D*<sub>*LE*</sub> (mm) | 0.99±0.08 | 1.07±0.11 | <0.001 |
|  |  |  | *W*<sub>*LE*</sub>(N.mm) | 1094.88±260.45 | 933.67±221.21 | <0.001 |
|  |  | *Plastic & irreversible* | *D*<sub>1</sub> (mm) | 1.82±0.24 | 1.99±0.23 | <0.001 |
|  |  |  | *D*<sub>*R*</sub>(mm) | 1.58±0.21 | 1.72±0.21 | <0.001 |
|  |  |  | *W*<sub>*D*</sub> (N.mm) | 3838.08±863.33 | 3374.73±735.07 | <0.001 |
|  |  |  | *W*<sub>*R*</sub> (N.mm) | 1680.36±389.83 | 1467.25±339.98 | <0.001 |

\* *P*−values were calculated using an unpaired two-sample t-test for normally distributed data and the Mann–Whitney U test otherwise.

AUROC, Harrell’s C-index, and AIC were evaluated for all FE-based descriptors (Supplementary Table B1), with complete results provided in Supplementary Table D1. Table 2 summarizes the selected FE-based descriptors showing significant improvements in discrimination or ΔAIC ≥ 10 relative to TH or FN aBMD, together with the results for aBMD and 3D-DXA-derived parameters.TH and FN aBMD showed similar AUROC values (0.763 and 0.757, respectively). Among 3D-DXA-derived parameters, Trab.vBMD achieved the highest AUROC (0.784), although none significantly outperformed aBMD. In contrast, *F*_*U*_ + *W*_*R*_ (AUROC = 0.797) and *F*_*LE*_ + *W*_*LE*_ (AUROC = 0.794) significantly outperformed both TH and FN aBMD (all P < 0.05), while *F*_*LE*_ + *D*_*LE*_ and *F*_*LE*_+ *W*_*R*_ showed significant improvements over TH aBMD only.

**Table 2.**
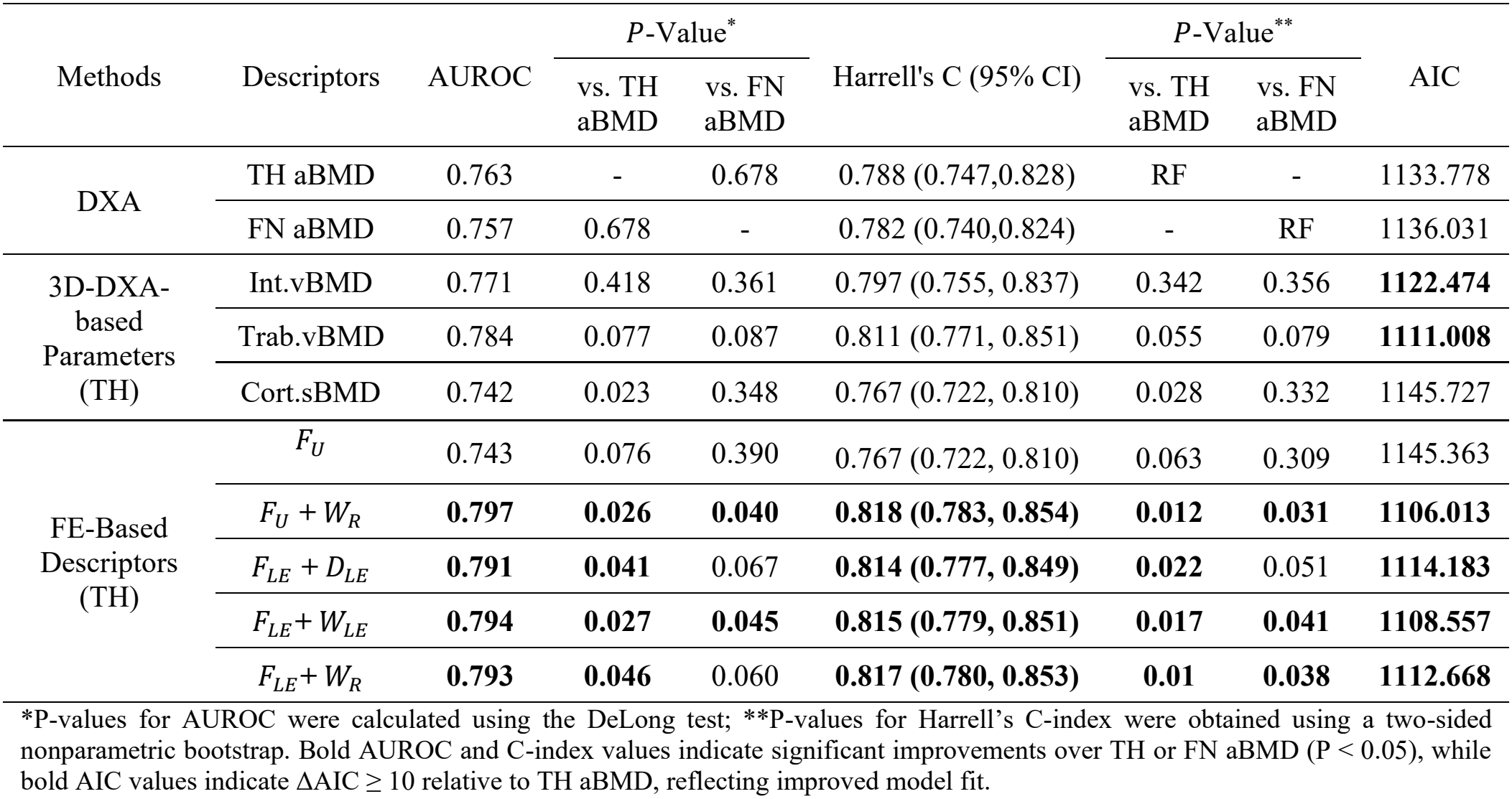
AUROC, Harrell’s C-index, and AIC comparison of DXA, 3D-DXA-based parameters, and FE-based descriptors.

| Methods | Descriptors | AUROC | *P*-Value<sup>\*</sup> |  | Harrell's C (95% CI) | *P*-Value<sup>\*\*</sup> |  | AIC |
|---|---|---|---|---|---|---|---|---|
|  |  |  | vs. TH aBMD | vs. FN aBMD |  | vs. TH aBMD | vs. FN aBMD |  |
| DXA | TH aBMD | 0.763 | - | 0.678 | 0.788 (0.747,0.828) | RF | - | 1133.778 |
|  | FN aBMD | 0.757 | 0.678 | - | 0.782 (0.740,0.824) | - | RF | 1136.031 |
| 3D-DXA-based Parameters (TH) | Int.vBMD | 0.771 | 0.418 | 0.361 | 0.797 (0.755, 0.837) | 0.342 | 0.356 | **1122.474** |
|  | Trab.vBMD | 0.784 | 0.077 | 0.087 | 0.811 (0.771, 0.851) | 0.055 | 0.079 | **1111.008** |
|  | Cort.sBMD | 0.742 | 0.023 | 0.348 | 0.767 (0.722, 0.810) | 0.028 | 0.332 | 1145.727 |
| FE-Based Descriptors (TH) | *F*<sub>*U*</sub> | 0.743 | 0.076 | 0.390 | 0.767 (0.722, 0.810) | 0.063 | 0.309 | 1145.363 |
|  | *F*<sub>*U*</sub> + *W*<sub>*R*</sub> | **0.797** | **0.026** | **0.040** | **0.818 (0.783, 0.854)** | **0.012** | **0.031** | **1106.013** |
|  | *F*<sub>*LE*</sub> + *D*<sub>*LE*</sub> | **0.791** | **0.041** | 0.067 | **0.814 (0.777, 0.849)** | **0.022** | 0.051 | **1114.183** |
|  | *F*<sub>*LE*</sub>+ *W*<sub>*LE*</sub> | **0.794** | **0.027** | **0.045** | **0.815 (0.779, 0.851)** | **0.017** | **0.041** | **1108.557** |
|  | *F*<sub>*LE*</sub>+ *W*<sub>*R*</sub> | **0.793** | **0.046** | 0.060 | **0.817 (0.780, 0.853)** | **0.01** | **0.038** | **1112.668** |

\*P-values for AUROC were calculated using the DeLong test; \*\*P-values for Harrell's C-index were obtained using a two-sided nonparametric bootstrap. Bold AUROC and C-index values indicate significant improvements over TH or FN aBMD (P < 0.05), while bold AIC values indicate ΔAIC ≥ 10 relative to TH aBMD, reflecting improved model fit.

Harrell’s C-index showed consistent findings. *F*_*U*_ + *W*_*R*_ (0.818), *F*_*LE*_ + *W*_*LE*_ (0.815), and *F*_*LE*_ + *W*_*R*_ (0.817) significantly outperformed both TH and FN aBMD (0.788 and 0.782, respectively; all P < 0.05). *F*_*LE*_ + *D*_*LE*_ also improved discrimination relative to TH aBMD (P = 0.022). AIC further supported improved model fit for these four FE-based combinations (1106.013–1114.183 vs. 1133.778 for TH aBMD; ΔAIC > 10), with similarly favorable AIC values for Trab.vBMD and Int.vBMD.

Net reclassification improvement (NRI) was evaluated for 3D-DXA-derived parameters and FE-based descriptors using TH and FN aBMD as reference models (Table 3), with complete results provided in Supplementary Table D2.

**Table 3.** NRI for DXA, 3D-DXA-based parameters, and FE-based descriptors vs. TH and FN aBMD.

| Methods | Descriptor | Vs TH aBMD as a reference |  | Vs FN aBMD as a reference |  |
|---|---|---|---|---|---|
|  |  | NRI* (95% CI) | *P*-Value<sup>\*\*</sup> | NRI* (95% CI) | *P*-Value<sup>\*\*</sup> |
| DXA | TH aBMD | Ref | - | - | - |
|  | FN aBMD | - | - | Ref | - |
| 3D-DXA-based Parameters (TH) | Int.vBMD | 0.027 (-0.049,0.172) | 0.484 | 0.106 (-0.006,0.226) | 0.072 |
|  | Trab.vBMD | 0.074 (-0.042, 0.221) | 0.241 | **0.159 (0.024, 0.311)** | **0.022** |
|  | Cort.sBMD | -0.038 (-0.085, 0.093) | 0.863 | 0.018 (-0.040, 0.186) | 0.439 |
| FE-Based Descriptors (TH) | *F<sub>U</sub>* | -0.001 (-0.059, 0.057) | 0.825 | 0.048 (-0.026, 0.164) | 0.320 |
|  | ***F<sub>U</sub> + W<sub>R</sub>*** | 0.114 (-0.002, 0.229) | 0.053 | **0.202 (0.075, 0.333)** | **p < 0.001** |
|  | ***F<sub>LE</sub> + D<sub>LE</sub>*** | 0.090 (-0.023, 0.191) | 0.129 | **0.161 (0.046, 0.307)** | **0.002** |
|  | ***F<sub>LE</sub>+ W<sub>LE</sub>*** | 0.093 (-0.004, 0.211) | 0.060 | **0.182 (0.071, 0.326)** | **0.001** |
|  | ***F<sub>LE</sub>+ W<sub>R</sub>*** | 0.123 (-0.016, 0.263) | 0.099 | **0.187 (0.047, 0.355)** | **0.002** |

\*NRI was calculated by adding each parameter to the reference model (TH or FN aBMD) to assess its added predictive value. \*\*P-values were obtained using two-sided bootstrap resampling. Bold descriptor names indicate selection based on AUROC performance (Table 2), while bold NRI values indicate statistically significant improvements (P < 0.05) relative to TH or FN aBMD.

Using FN aBMD as the reference model, several parameters demonstrated significant improvements in risk reclassification. Among the 3D-DXA-based measurements, only Trab.vBMD showed a statistically significant improvement versus FN aBMD (NRI = 0.159; 95% CI, 0.024–0.311; P = 0.022), whereas Int.vBMD and Cort.sBMD were not statistically significant.

Notably, multiple FE-based descriptors yielded significant improvements in reclassification performance relative to FN aBMD. In particular, *F*_*U*_ + *W*_*R*_ showed the highest NRI (0.202; 95% CI, 0.075–0.333; P < 0.001), followed by *F*_*LE*_ + *D*_*LE*_ (P = 0.002), *F*_*LE*_+ *W*_*LE*_ (P = 0.001), and *F*_*LE*_+ *W*_*R*_ (P = 0.002), indicating improved classification of individuals into appropriate risk categories when combined with FN aBMD.

In comparison, when TH aBMD was used as the reference model, improvements were more moderate. FE-based descriptors such as *F*_*U*_ + *W*_*R*_ (P = 0.053) and *F*_*LE*_ + *W*_*LE*_ (P = 0.060) showed positive NRI values with borderline statistical significance, suggesting a consistent trend toward improved risk classification relative to TH aBMD.

To complement the NRI analysis (Table 3), IDI and cNRI were evaluated to further assess predictive performance and risk reclassification beyond aBMD, without reliance on predefined risk categories. Complete results are provided in Supplementary Table D3. FE-based combined descriptors consistently showed positive IDI and cNRI values, with significant improvements relative to both TH and FN aBMD. Among these, F_U_ + W_R_ demonstrated the highest IDI (0.036 and 0.042) and cNRI (0.570 and 0.459) relative to TH and FN aBMD, respectively (all P < 0.001). Similar improvements were observed for the other selected FE-based combinations, supporting their added predictive value beyond aBMD.

HRs for the selected FE-based descriptors (Table 2) are presented in Table 4, with complete results in Supplementary Table D4. TH and FN aBMD were significantly associated with hip fracture risk (HR = 2.696 and 2.746, respectively; both P < 0.001). All 3D-DXA-derived parameters also showed significant associations, with Trab.vBMD yielding the highest HR (3.268; 95% CI: 2.702–4.035). Among FE-based descriptors, *F*_*U*_ *+ W*_*R*_ (HR = 3.577; 95% CI: 2.821–4.597) and *F*_*LE*_*+ W*_*R*_ (HR = 3.541; 95% CI: 2.797–4.515) showed the highest HRs, followed by *F*_*LE*_*+ W*_*LE*_ (HR = 3.330; 95% CI: 2.623–4.281; all P < 0.001). For these FE-based descriptors, the 95% confidence intervals were shifted toward higher HR ranges compared with both TH aBMD and FN aBMD, while still overlapping with those of aBMD, further supporting their stronger association with fracture risk.

**Table 4.** HRs with 95% CI per one SD change for DXA, 3D-DXA-based parameters, and FE-based descriptors.

|  |  | HR (95% CI) | *P*-Value<sup>\*\*</sup> |
|---|---|---|---|
| DXA | TH aBMD | 2.696 (2.289, 3.223) | <0.001 |
|  | FN aBMD | 2.746 (2.299, 3.316) | <0.001 |
| 3D-DXA-based Parameters (TH) | Int.vBMD | **2.978 (2.485, 3.638)** | <0.001 |
|  | Trab.vBMD | **3.268 (2.702, 4.035)** | <0.001 |
|  | Cort.sBMD | 2.515 (2.121, 3.026) | <0.001 |
| FE-Based Descriptors (TH**)** | *F<sub>U</sub>* | 2.693 (2.228, 3.291) | <0.001 |
|  | ***F<sub>U</sub> + W<sub>R</sub>*** | **3.577 (2.821, 4.597)** | <0.001 |
|  | ***F<sub>LE</sub> + D<sub>LE</sub>*** | 2.421 (1.987, 3.028) | <0.001 |
|  | ***F<sub>LE</sub>+ W<sub>LE</sub>*** | **3.330 (2.623, 4.281)** | <0.001 |
|  | ***F<sub>LE</sub>+ W<sub>R</sub>*** | **3.541 (2.797, 4.515)** | <0.001 |

\* HRs are reported per 1-SD change in each predictor. \*\* P-Values were obtained via nonparametric bootstrap resampling; Bold descriptors reflect AUROC-based selection (Table 2), and bold HR values indicate estimates greater than TH aBMD.

The ROC curves for the FE-based descriptors (*F*_*U*_ + *W*_*R*_ and *F*_*LE*_ + *W*_*LE*_), which showed the highest AUROC values with statistically significant improvement compared with both TH aBMD and FN aBMD, are presented in Fig. 2.

**Fig. 2.**
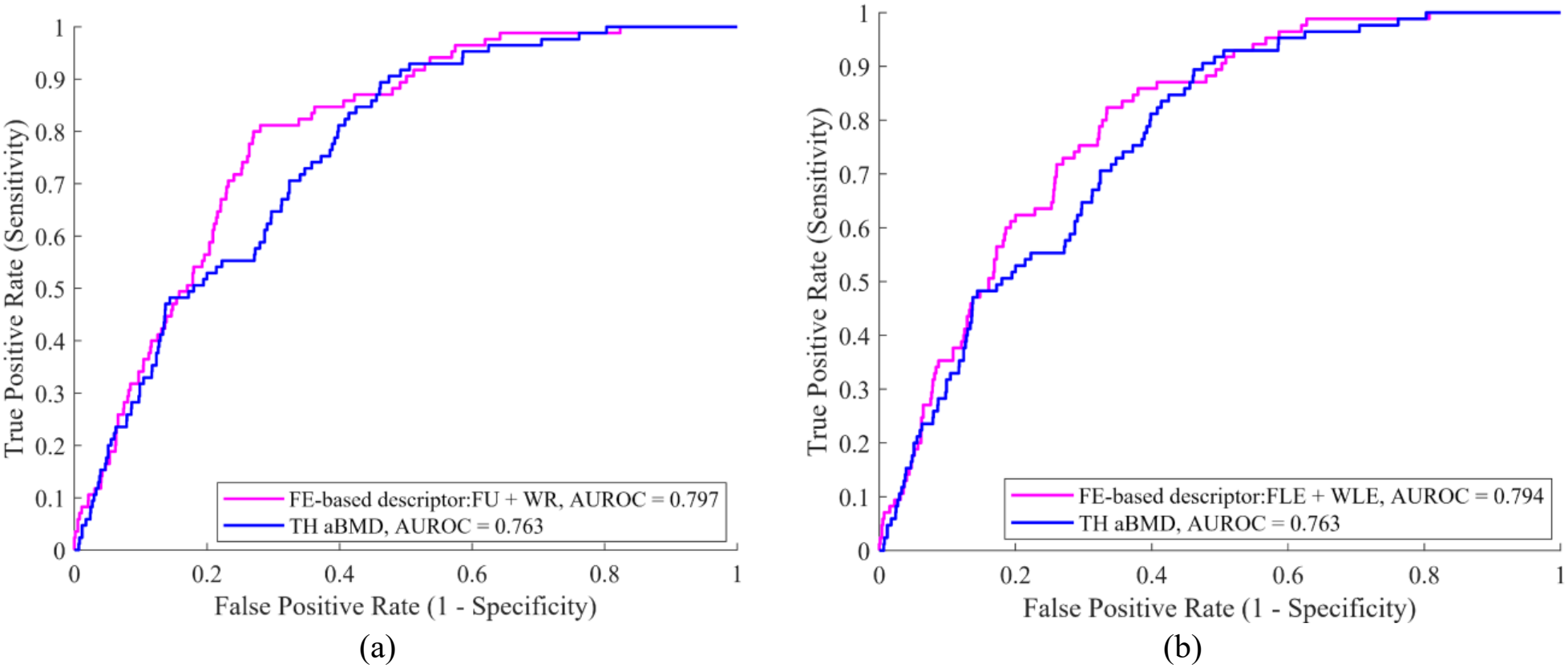
ROC curves comparing the FE-based descriptor: *F*_*U*_ + *W*_*R*_ (a), and *F*_*LE*_ + *W*_*LE*_ (b) with TH aBMD

## Discussion

To our knowledge, this is the first study to demonstrate that global FE-based mechanical descriptors derived from standard hip DXA scans outperform aBMD in hip fracture prediction within a prospective, population-based cohort of women. Notably, TH and FN aBMD already showed relatively high discrimination (AUROC = 0.763 and 0.757, respectively) compared with previous studies [6, 17, 19], making further improvements more challenging. Nevertheless, more than two-thirds of women who experienced hip fractures were not classified as osteoporotic based on TH aBMD, with only a minority meeting the FN aBMD osteoporosis threshold. These findings highlight the limitations of relying on aBMD alone and suggest that FE-based mechanical descriptors may help identify women with mechanically vulnerable proximal femurs despite non-osteoporotic aBMD values.

FE-based descriptors showed consistent improvements across complementary statistical analyses, including discrimination (AUROC and Harrell’s C-index), model fit (AIC), reclassification (NRI, IDI, and cNRI), and risk association (HR). In particular, the combined descriptors *F*_*U*_ + *W*_*R*_, *F*_*LE*_ +

*D*_*LE*_, *F*_*LE*_+ *W*_*LE*_, and *F*_*LE*_+ *W*_*R*_ demonstrated improved performance relative to aBMD across several metrics (Tables 2–4), whereas 3D-DXA-derived parameters generally showed comparable discrimination to aBMD. By combining strength with deformation and energy-related measures, these FE-based descriptors characterize complementary aspects of femoral mechanical behavior under subject-specific 3D geometry, density distribution, and simulated side-fall loading that are not captured by aBMD alone.

Previous QCT-based FE studies by Orwoll et al. [7], Amin et al. [8], and Kopperdahl et al. [33] reported fracture discrimination comparable to aBMD using femoral strength and load-to-strength ratios. In contrast, Adams et al. [4] demonstrated improved fracture prediction using FE-derived strength within the Biomechanical Computed Tomography (BCT) framework, while Keaveny et al. [34] reported further improvements by combining strength with clinical variables. Building on these findings, the present study extends FE-based fracture prediction to a DXA-based framework by combining strength with deformation- and energy-related descriptors to characterize femoral mechanical behavior beyond strength alone. Moreover, the higher cost and radiation exposure associated with QCT support the development of more accessible DXA-based FE approaches.

Previous studies using DXA-based 2D FE models reported marginal or inconsistent improvements over aBMD in fracture risk prediction using femoral strength, load-to-strength ratios, or Bone Strain Index [13, 35]. Their planar representation limits the characterization of subject-specific 3D femoral geometry, loading conditions, and mechanical response. In contrast, the present 3D-DXA-based FE approach demonstrated statistically significant improvements over aBMD across several measures of predictive performance.

Previous 3D-DXA-based FE studies by Grassi et al. [6, 17] demonstrated improved fracture discrimination using weight-normalized femoral strength under multiple simulated side-fall configurations in the MrOS and OSTPRE cohorts, particularly relative to FN aBMD in OSTPRE. However, these studies employed linear elastic FE models with a surface strain-based failure criterion [36], whereas the present study incorporates nonlinear material behavior to characterize the volumetric inelastic response of the femur. Although differences between cohorts prevent direct comparisons of predictive performance, previous studies have demonstrated improved fracture discrimination using nonlinear FE models [5, 19]. Furthermore, while previous 3D-DXA-based FE studies primarily focused on femoral strength, our earlier study [19] demonstrated the additional discriminative value of nonlinear energy-related descriptors, which is further supported by the present findings in an independent prospective cohort.

Another relevant 3D-DXA-based FE study by Gugler et al. [23] investigated fracture prediction in the prospective Geneva Retirees Cohort, evaluating femoral strength under simulated fall conditions alongside 3D-DXA-derived parameters. Consistent with our findings, femoral strength showed a fracture risk association comparable to TH aBMD, whereas Trab.vBMD showed the strongest association among the evaluated FE strength and 3D-DXA-derived parameters, in agreement with Iki et al. [37] in the JPOS cohort. Gugler et al. [23] also reported numerically higher Harrell’s C-index values for FE strength and vBMD than for aBMD, without statistically significant differences, consistent with our findings for Int.vBMD and Trab.vBMD (Table 2). However, methodological and cohort-related differences should be considered. The voxel-based FE approach of Gugler et al. [23] does not explicitly distinguish cortical and trabecular compartments [38], whereas our compartment-specific structural mesh represents their distinct morphology and mechanical properties. Furthermore, their cohort included only five hip fractures, with a shorter follow-up period, compared with 85 hip fractures and up to 20 years of follow-up in the present study. Gugler et al. [23] also reported that strength-based thresholds improved fracture identification but increased the classification of non-fracture individuals as high risk, reducing specificity. These findings suggest the need to investigate thresholds combining strength with deformation- and energy-related descriptors. The present prospective results, together with our previous retrospective study [19] (64 fractures and 64 controls), provide a basis for evaluating such thresholds in future studies.

From a computational perspective, the proposed framework requires approximately 1.5 min for 3D reconstruction (Intel Core i7-4790K CPU) and 10 min for FE simulation (8 CPU cores) per subject, supporting its potential clinical applicability.

Despite its advantages, the proposed approach has several limitations. The relatively limited number of hip fractures (85 cases among 1,636 women) may affect the statistical power to detect improvements over aBMD [37]. Internal cross-validation, bootstrap resampling, and penalized regression were used to reduce overfitting and improve model robustness. Although the findings are consistent with our previous independent Spanish case-control study [19], validation in larger and more diverse cohorts is needed, particularly given the inclusion of Japanese women only and potential population-related differences in femoral geometry and density distribution. Furthermore, the elastic– plastic constitutive model approximates bone failure mechanisms, and more advanced macroscopic or multiscale failure models [39–41] warrant investigation. Finally, global FE-based descriptors were evaluated, as fracture location was unavailable. Although local descriptors may improve site-specific fracture prediction [22], global descriptors offer greater computational efficiency and robustness.

## Conclusion

This is the first study demonstrating that global mechanical descriptors obtained from standard hip DXA scans outperform aBMD in predicting incident hip fractures in a population-based, prospectively followed cohort of women. By incorporating mechanical descriptors beyond bone density alone, these descriptors may provide a more comprehensive characterization of skeletal fragility. These findings highlight the potential of integrating 3D-DXA and FE modelling into existing DXA-based clinical workflows to enhance fracture risk assessment.

## Declarations

## Acknowledgements

This study was funded by the Spanish Government (CPP2021-008574, CPP2023-010970) and AGAUR (2023 BP 00224). The baseline survey of JPOS study was supported by the Japan Milk Promotion Board and the Japan Dairy Council. The follow-up surveys were supported by Grants-in-aid for Scientific Research (B#10470114 [1998–2000]; B#14370147 [2002–2003]; B#18390201 [2006–2008]; C#18590619 [2006-2007]; B#23390180 [2011–2013]; C#23500852[2011-2013]; #23659362 [2011–2012]; #23657176 [2011–2012]; and B#24390505 [2012–2014], A#15H02526 [2015–2019]; B#15H05102 [2015–2017]; B#15H04789 [2015–2017]; #16K15384; and #16K19263 [2016–2017]) from the Japanese Society for the Promotion of Science, a grant in 2000–2002 from the Research Society for Metabolic Bone Diseases, Japan, a Grant-in-Aid to Study Milk Nutrition (2006, 2008, 2015, 2016) from the Japan Dairy Association, and a Grant-in-Aid (2015) from the Univers Foundation.

The cohort was conducted by the JPOS Study Group, comprising Masayuki Iki (Chairman of the Study Group), Matsumoto, Toshio (Professor Emeritus Tokushima University) along with the co-authors. We are deeply grateful to the residents and the public health staff of Sanuki City, Nishiaizu Town, Miyakojima City, Joetsu City, and Memuro Town for their valuable cooperation and support of these surveys. We also would like to express our sincere gratitude to those from Toyo Medic, Osaka, Japan and to Mr. Yasuhiko Takegahara, President of Take Medical Service, for his invaluable technical support in this study. In addition, we would like to express our deep thanks to Dr. Kiichi Nonaka, Scientific Support Manager, TOYO MEDIC Co., Ltd., for organizing and making progress ahead about the present study.

## Author contributions

**Elham Alizadeh:** Writing, review & editing; study design; methodology and implementation; analysis and interpretation of the results. **Junko Tamaki:** Principal investigator of the JPOS Study; supervised the JPOS surveys; interpreted the JPOS data; reviewed the manuscript. **Abdollah Moslemi:** Review & editing; methodology; analysis. **Takahiro Tachiki:** Acquisition of follow-up survey JPOS data; reviewed the manuscript. **Yuho Sato:** Acquisition of follow-up survey JPOS data; reviewed the manuscript. **Masami Hamada:** Acquisition of fracture event JPOS data; reviewed the manuscript. **Kouji Tsuda:** Interpreted the JPOS data; reviewed the manuscript. **Asako Kudo:** Interpreted the JPOS data; reviewed the manuscript. **Kuniyasu Kamiya:** Acquisition of follow-up survey JPOS data; reviewed the manuscript. **Nami Imai:** Acquisition of follow-up survey JPOS data; reviewed the manuscript. **Akemi Nitta:** Acquisition of follow-up survey JPOS data; reviewed the manuscript. **Kumiko Ohara:** Interpreted the JPOS data; reviewed the manuscript. **Naoyuki Takashima:** Interpreted the JPOS data; reviewed the manuscript. **Jun Kitagawa:** Acquisition of follow-up survey JPOS data; reviewed the manuscript. **Katsuyasu Kouda:** Acquisition of follow-up survey JPOS data; reviewed the manuscript. **Akemi Morita:** Acquisition of follow-up survey JPOS data; reviewed the manuscript. **Kazuhiro Uenishi:** Acquisition of follow-up survey JPOS data; reviewed the manuscript. **Etsuko Kajita:** Acquisition of fracture event JPOS data; reviewed the manuscript. **Sadanobu Kagamimori:** Conceptualized and conducted the JPOS Study; reviewed the manuscript. **Ludovic Humbert:** Study design; methodology; interpretation of the results; review & editing. **Jerome Noailly:** Study design; methodology; interpretation of the results; review & editing; funding. **Masayuki Iki:** Chairman of the JPOS Study Group; conceptualized the JPOS Study; conducted the JPOS data surveys; interpreted the data; reviewed the manuscript.

## Ethical approval

The JPOS baseline and follow-up surveys were conducted in accordance with the Declaration of Helsinki and approved by the Ethics Committees of Kindai University Faculty of Medicine (no. 30-133), Osaka Medical College (no. 1757-02), and Osaka Medical and Pharmaceutical University (no. 2021-070-9, May 30, 2024). Written informed consent was obtained from all participants before each survey. Secondary data use for the present study was approved by the Ethics Committee of Osaka Medical and Pharmaceutical University (no. 2023-131-1, October 10, 2023).

## Data availability statement

The main results supporting this study’s findings are available from the corresponding author on request. The data are not publicly available due to privacy or ethical restrictions.

## Informed consent

Informed consent was obtained from all individual participants included in the study. Written informed consent was obtained before each survey.

