## Supplementary Materials for "3D-DXA-based Finite Element Models Improve Hip Fracture Prediction versus aBMD in the Japanese Population-Based Osteoporosis (JPOS) Cohort Study"

**Elham Alizadeh**[1], **Junko Tamaki**[2], **Abdollah Moslemi**[1], **Takahiro Tachiki**[3], **Yuho Sato**[4], **Masami Hamada**[5], **Kouji Tsuda**[2], **Asako Kudo**[2], **Kuniyasu Kamiya**[6], **Nami Imai**[7], **Akemi Nitta**[8], **Kumiko Ohara**[9], **Naoyuki Takashima**[9], **Jun Kitagawa**[10], **Katsuyasu Kouda**[8], **Akemi Morita**[11], **Kazuhiro Uenishi**[12], **Etsuko Kajita**[13], **Sadanobu Kagamimori**[14], **Ludovic Humbert**[15], **Jérôme Noailly**[1], **Masayuki Iki**[8] ([1]Pompeu Fabra University, Spain; [2] Osaka Medical and Pharmaceutical University, Japan; [3] University of Kochi, Japan; [4] Jin-ai University, Japan; [5] Josai International University, Japan; [6] Kobe City College of Nursing, Japan; [7] Nihon Pharmaceutical University, Japan; [8] Kansai Medical University, Japan; [9] Kyoto Prefectural University of Medicine, Japan; [10] Kitasato University, Japan; [11] Tottori University, Japan; [12] Japan Nutrition University, Japan; [13] Nagoya University, Japan; [14] University of Toyama, Japan; [15] 3D-Shaper Medical, Spain)

### Supplementary A

Fig. A1 illustrates a schematic representation of the FE-based mechanical descriptors derived from the femoral force–displacement response.

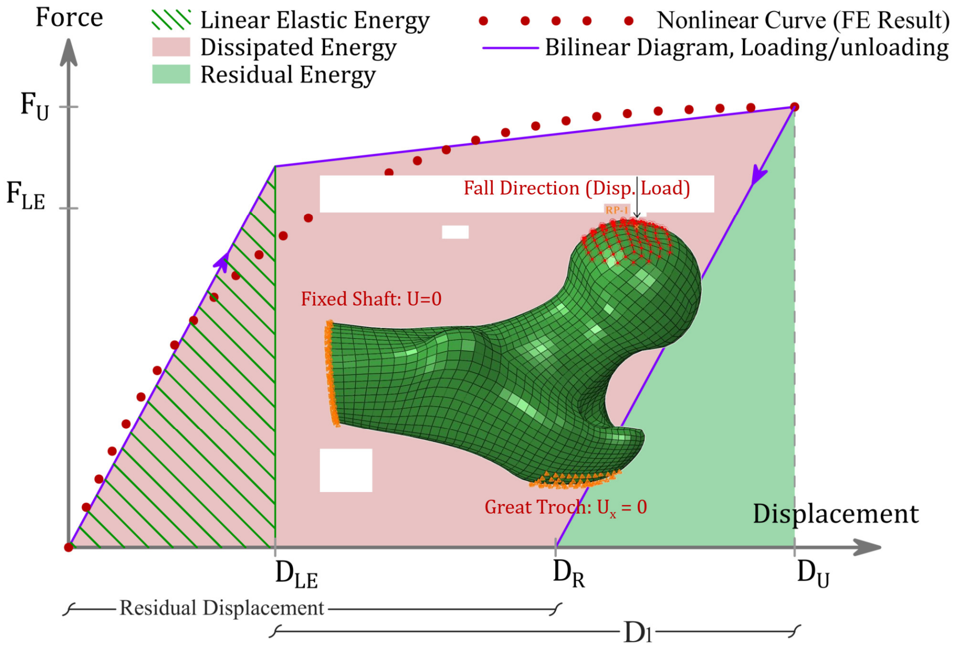


Fig. A1. Schematic overview of FE-based mechanical descriptors derived from the force–displacement response of the femur.

### Supplementary B

FE-based descriptors derived from nonlinear mechanical responses, piecewise-linear analyses, and their combinations is presented in Table B1.

Table B1. FE-based descriptors obtained from the nonlinear response, piecewise-linear analyses, and combined descriptors.

| Analysis | Mechanical phase | Descriptor type | Descriptors |
|---|---|---|---|
| Nonlinear FE response | Global | *Peak/failure* | Strength ($F_U$), Displacement ($D_U$), Nonlinear Energy ($W_U$) |
| Piecewise-linear analysis | Linear Elastic regime | *Elastic* | Linear Elastic Force ($F_{LE}$), Linear Elastic Displacement ($D_{LE}$), Linear elastic energy ($W_{LE}$) |
| | Post-yield / plastic regime | *Plastic & irreversible* | Nonlinear Deformation ($D_1$), Residual Displacement ($D_R$), Dissipated Energy ($W_D$), Residual Energy ($W_R$) |
| Combined Descriptors | Maximum load + deformation/energy | *Strength-Based* | $F_U$+ $D_U$, $F_U$+ $W_U$, $F_U$+ $D_1$, $F_U$+ $D_R$, $F_U$+ $W_D$, $F_U$+ $W_R$ |
| | | *Linear Elastic Force- Based* | $F_{LE}$ + $D_{LE}$, $F_{LE}$+ $W_{LE}$, $F_{LE}$ + $D_1$, $F_{LE}$+ $D_R$, $F_{LE}$ + $W_D$, $F_{LE}$+ $W_R$ |

### Supplementary C. Detailed Statistical Methodology

The predictive performance of the 3D-DXA-based FE mechanical descriptors was evaluated through group comparisons, fracture discrimination, and Cox survival analyses. The survival analyses comprised four complementary assessments: discrimination, model fit, reclassification, and associations with fracture risk. The overall statistical workflow is illustrated in Supplementary Fig. C1.

#### C1. Group Differences

Differences between subjects who experienced hip fractures and controls were assessed using unpaired t-tests for normally distributed variables or Mann–Whitney U tests for non-normally distributed variables. Statistical significance was assessed using the corresponding *p*-values.

#### C2. Fracture Discrimination

The ability of individual FE mechanical descriptors to discriminate between fracture and control subjects was evaluated using logistic regression and receiver operating characteristic analysis. Discriminative performance was quantified using the area under the receiver operating characteristic curve (AUROC).

Multivariable logistic regression models were constructed to evaluate combinations of mechanical descriptors for hip fracture discrimination. Model performance was assessed using leave-one-out cross-validation (LOOCV), and differences between AUROC values were evaluated using the DeLong test.

#### C3. Cox Survival Modelling

To extend the evaluation to time-to-event outcomes, Cox proportional hazards models were used to incorporate fracture timing and censoring information. The survival analyses included discrimination using Harrell's C-index, model fit using Akaike's Information Criterion (AIC), reclassification using NRI, IDI, and cNRI, and associations with fracture risk using ridge-penalized Cox regression.

##### C3.1. Discrimination: Harrell's C-index

Model discrimination was assessed within a survival analysis through Cox models. Harrell's C-index [27] was used to evaluate the discriminative ability of each Cox model to correctly rank individuals, by assigning higher risks to subjects who experienced fractures earlier compared to those who fractured later or to those who remained fracture-free. C-indices were computed based on subject pairs, accounting for time-to-event information and censoring. They were derived from feature (e.g., aBMD, Strength, …)-based risk scores, and paired differences (ΔC) were calculated to assess improvements in discrimination relative to reference feature.

Nonparametric bootstrap resampling was used to estimate 95% CIs and two-sided *p*-values for ΔC. The bootstrap procedure preserved the paired comparison between models by evaluating both models within the same resampled subjects.

##### C3.2. Model Fit: Akaike's Information Criterion

Model fit was assessed using Akaike's Information Criterion (AIC), which was used to compare the relative goodness-of-fit of the Cox models while accounting for model complexity. Lower AIC values indicate a more favorable balance between explanatory power and model parsimony. Differences in AIC (ΔAIC), in different descriptors, were used to compare competing models, with values $< 2$ indicating comparable performance, values of 4–7 indicating meaningful differences, and values $> 10$ indicating substantial improvement of the model with the lower AIC [28].

##### C3.3. Reclassification: NRI, IDI, and cNRI

Beyond discrimination and model fit, the additional predictive value of FE-based descriptors was evaluated using categorical net reclassification improvement (NRI), integrated discrimination improvement (IDI), and continuous NRI (cNRI).

**Categorical net reclassification improvement (NRI)**

Categorical NRI [29] was calculated by comparing reference Cox models based on total hip (TH) or femoral neck (FN) areal bone mineral density (aBMD) with extended models incorporating FE-based descriptors.

Participants were stratified into low-, intermediate-, and high-risk categories according to their predicted fracture probabilities. Risk thresholds were defined by mapping standard T-score thresholds to corresponding aBMD values using NHANES III young-adult female reference data for the proximal femur [31]. These aBMD thresholds were subsequently translated into predicted fracture-risk thresholds using the reference Cox model.

The same risk thresholds were applied to both reference and extended models to ensure consistent risk classification. Reclassification analyses were performed using the full available follow-up period.

**Integrated discrimination improvement (IDI)**

Integrated discrimination improvement (IDI) [29] was calculated to quantify changes in risk discrimination between the extended and reference models. Risk discrimination was defined as the difference between the mean predicted fracture risks in fracture and control groups. IDI was calculated as the difference in this discrimination measure between the extended and reference models.

As a category-independent measure, IDI did not rely on predefined risk thresholds and directly quantified improvements in the separation of predicted risks between fracture and control subjects.

**Continuous net reclassification improvement (cNRI)**

Continuous NRI (cNRI), also referred to as category-free NRI [29,30], was calculated to assess improvements in predicted fracture risk without predefined risk categories.

The analysis was based on the direction of change in predicted fracture risk between reference and extended models. An upward change in predicted risk for fracture subjects and a downward change for controls were considered favorable reclassification [29,30].

**Statistical inference**

For all three reclassification measures (categorical NRI, IDI, and cNRI), 95% confidence intervals (CIs) and *p*-values were estimated using nonparametric bootstrap resampling.

#### C3.4. Association with Fracture Risk: Ridge-Penalized Cox Regression

To quantify the strength of association between individual descriptors, their combinations, and fracture risk, hazard ratios (HRs) were estimated using ridge-penalized Cox proportional hazards models [32].

Time to first hip fracture was defined as the outcome, and control subjects were treated as censored at their last available follow-up.

To enable comparisons between variables expressed in different physical units, all predictors were standardized by mean-centering and scaling to unit variance. HRs were reported per one standard deviation (SD) change in each predictor.

Ridge penalization was used to mitigate potential multicollinearity among FE-based descriptors and improve model stability given the limited number of fracture events.

The penalization parameter ($\lambda$) was tuned using 10-fold cross-validation. The optimal $\lambda$ was selected by maximizing the cross-validated partial log-likelihood. The largest optimal $\lambda$ value was subsequently applied across all models.

HRs with 95% CIs and *p*-values were estimated using nonparametric bootstrap resampling.

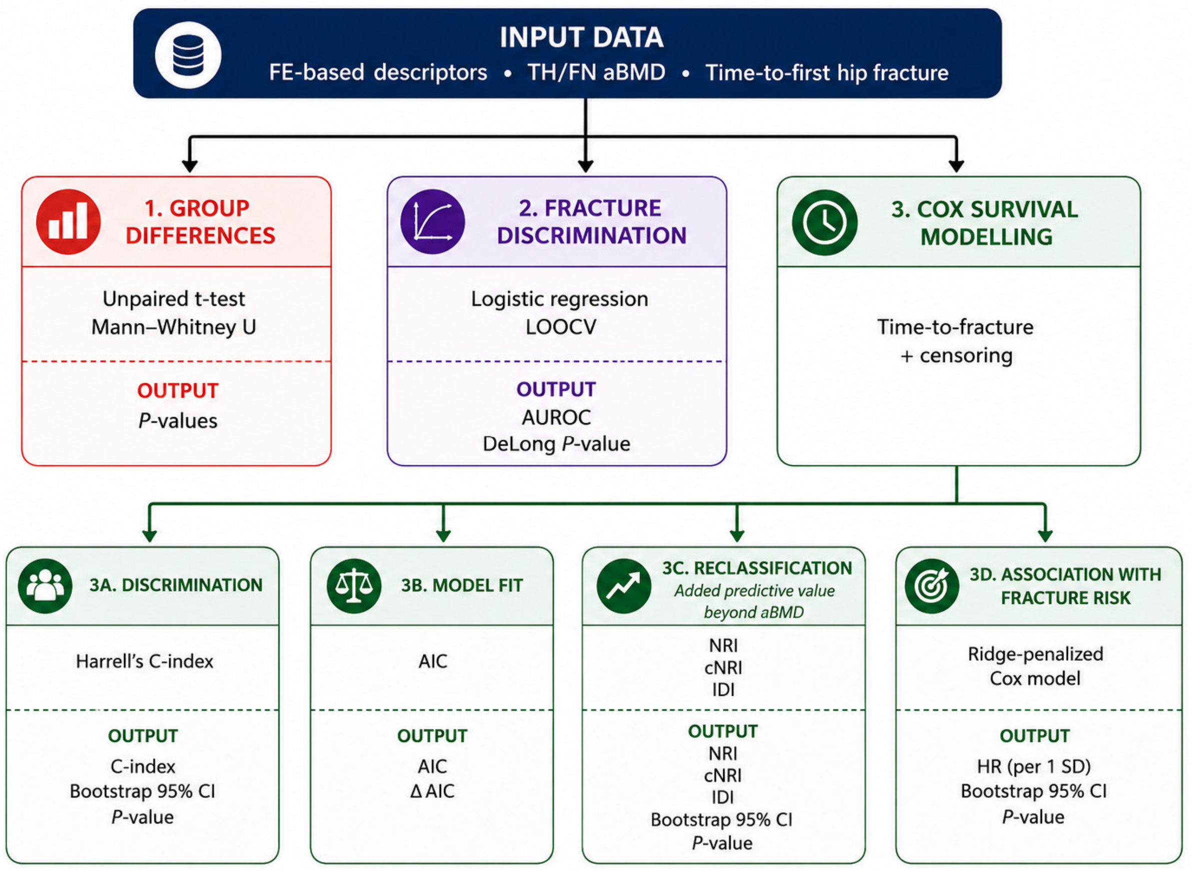


Fig. C1. Overview of the statistical analysis workflow.

### Supplementary D

The comprehensive performance of DXA, 3D-DXA–based measurements, and all FE-based descriptors was evaluated using AUROC, Harrell's C-index, and model fit criteria (AIC), with results summarized in Table D1.

Table D1. AUROC, Harrell's C-index, and AIC values for DXA, 3D-DXA-based measurements, and all FE-based descriptors evaluated for hip fracture prediction (see Table 1 for descriptor definitions).

| Methods | | | Descriptors | AUROC | $P$-Value* | | Harrell's C (95% CI) | $P$-Value** | | AIC |
|---|---|---|---|---|---|---|---|---|---|---|
| | | | | | vs. TH aBMD | vs. FN aBMD | | vs. TH aBMD | vs. FN aBMD | |
| **DXA** | | | TH aBMD | 0.763 | - | 0.678 | 0.788 (0.747,0.828) | RF | - | 1133.778 |
| | | | FN aBMD | 0.757 | 0.678 | - | 0.782 (0.740,0.824) | - | RF | 1136.031 |
| **3D-DXA–Based Parameters (TH)** | | | Int.vBMD | 0.771 | 0.418 | 0.361 | 0.797 (0.755, 0.837) | 0.342 | 0.356 | **1122.474** |
| | | | Trab.vBMD | 0.784 | 0.077 | 0.087 | 0.811 (0.771, 0.851) | 0.055 | 0.079 | **1111.008** |
| | | | Cort.sBMD | 0.742 | 0.023 | 0.348 | 0.767 (0.722, 0.810) | 0.028 | 0.332 | 1145.727 |
| **FE-Based Descriptors (TH)** | **Nonlinear FE Analysis** | | $F_U$ | 0.743 | 0.076 | 0.390 | 0.767 (0.722, 0.810) | 0.063 | 0.309 | 1145.363 |
| | | | $D_U$ | 0.721 | 0.162 | 0.259 | 0.727 (0.676, 0.777) | 0.022 | 0.065 | 1171.211 |
| | | | $W_U$ | 0.656 | <0.001 | <0.001 | 0.676 (0.615, 0.734) | <0.001 | <0.001 | 1191.002 |
| | **Piecewise Linear Analysis** | *Elastic* | $F_{LE}$ | 0.751 | 0.252 | 0.700 | 0.775 (0.732, 0.816) | 0.216 | 0.625 | 1141.624 |
| | | | $D_{LE}$ | 0.721 | 0.237 | 0.326 | 0.720 (0.665, 0.775) | 0.046 | 0.071 | 1170.550 |
| | | | $W_{LE}$ | 0.684 | <0.001 | 0.001 | 0.707 (0.651, 0.762) | <0.001 | <0.001 | 1179.856 |
| | | *Plastic & irreversible* | $D_1$ | 0.709 | 0.058 | 0.111 | 0.718 (0.667, 0.767) | 0.009 | 0.031 | 1179.127 |
| | | | $D_R$ | 0.695 | 0.0193 | 0.042 | 0.704 (0.654, 0.752) | 0.004 | 0.011 | 1184.815 |
| | | | $W_D$ | 0.658 | <0.001 | <0.001 | 0.676 (0.618, 0.734) | <0.001 | <0.001 | 1191.553 |
| | | | $W_R$ | 0.659 | <0.001 | <0.001 | 0.681 (0.621, 0.739) | <0.001 | <0.001 | 1189.069 |
| | **Combined Descriptors** | *Strength-Based* | $F_U + D_U$ | 0.771 | 0.435 | 0.388 | 0.800 (0.763, 0.837) | 0.234 | 0.273 | 1130.037 |
| | | | $F_U + W_U$ | 0.768 | 0.636 | 0.500 | 0.797 (0.760, 0.834) | 0.356 | 0.338 | 1128.135 |
| | | | $F_U + D_1$ | 0.760 | 0.766 | 0.861 | 0.791 (0.753, 0.829) | 0.719 | 0.564 | 1135.692 |
| | | | $F_U + D_R$ | 0.753 | 0.330 | 0.802 | 0.785 (0.748, 0.824) | 0.837 | 0.832 | 1138.577 |
| | | | $F_U + W_D$ | 0.753 | 0.347 | 0.778 | 0.784 (0.746, 0.823) | 0.755 | 0.884 | 1136.424 |
| | | | $\boldsymbol{F_U + W_R}$ | **0.797** | **0.026** | **0.040** | **0.818 (0.783, 0.854)** | **0.012** | **0.031** | **1106.013** |
| | | *Linear Elastic Force-Based* | $\boldsymbol{F_{LE} + D_{LE}}$ | **0.791** | **0.041** | 0.067 | **0.814 (0.777, 0.849)** | **0.022** | 0.051 | **1114.183** |
| | | | $\boldsymbol{F_{LE} + W_{LE}}$ | **0.794** | **0.027** | **0.045** | **0.815 (0.779, 0.851)** | **0.017** | **0.041** | **1108.557** |
| | | | $F_{LE} + D_1$ | 0.763 | 0.982 | 0.703 | 0.794 (0.756, 0.832) | 0.487 | 0.435 | 1134.217 |
| | | | $F_{LE} + D_R$ | 0.759 | 0.659 | 0.927 | 0.790 (0.752, 0.828) | 0.764 | 0.595 | 1135.972 |
| | | | $F_{LE} + W_D$ | 0.762 | 0.953 | 0.752 | 0.792 (0.754, 0.830) | 0.627 | 0.498 | 1132.464 |
| | | | $\boldsymbol{F_{LE} + W_R}$ | **0.793** | **0.046** | 0.060 | **0.817 (0.780, 0.853)** | **0.01** | **0.038** | **1112.668** |

*P-Values calculated between AUROCs using the DeLong method; ** P-value for Harrell's C derived from two-sided nonparametric bootstrap of ΔC (C-index difference); bold descriptor names indicate selection based on AUROC performance (Table 2), whereas bold AUROC and Harrell C-index denote statistically significant improvement in predictive performance relative to TH aBMD or FN aBMD. Bold AIC values indicate ΔAIC ≥10 vs. TH aBMD, consistent with strong evidence of improved model fit.

To assess the added predictive value beyond DXA, net reclassification improvement (NRI) analyses were performed for 3D-DXA–based parameters and FE-based descriptors, using TH aBMD and FN aBMD as reference models (Table D2).

Table D2. NRI for DXA, 3D-DXA–based parameters, and FE-based descriptors vs. TH and FN aBMD

| Methods | | Descriptor | Vs TH aBMD as a reference | | Vs FN aBMD as a reference | |
|---|---|---|---|---|---|---|
| | | | NRI* (95% CI) | *P*-Value** | NRI* (95% CI) | *P*-Value** |
| **DXA** | | TH aBMD | Ref | - | - | - |
| | | FN aBMD | - | - | Ref | - |
| **3D-DXA–Based Parameters (TH)** | | Int.vBMD | 0.027 (-0.049,0.172) | 0.484 | 0.106 (-0.006,0.226) | 0.072 |
| | | Trab.vBMD | 0.074 (-0.042, 0.221) | 0.241 | **0.159 (0.024, 0.311)** | **0.022** |
| | | Cort.sBMD | -0.038 (-0.085, 0.093) | 0.863 | 0.018 (-0.040, 0.186) | 0.439 |
| **FE-Based Descriptors (TH)** – **Nonlinear FE Analysis** | | $F_U$ | -0.001 (-0.059, 0.057) | 0.825 | 0.048 (-0.026, 0.164) | 0.320 |
| | | $D_U$ | 0.009 (-0.073, 0.113) | 0.856 | 0.081 (-0.014, 0.218) | 0.096 |
| | | $W_U$ | -0.006 (-0.059, 0.117) | 0.834 | 0.031 (-0.039, 0.102) | 0.596 |
| **Piecewise Linear Analysis** | *Elastic* | $F_{LE}$ | -0.005 (-0.050, 0.058) | 0.722 | 0.085 (-0.027, 0.173) | 0.242 |
| | | $D_{LE}$ | 0.072 (-0.042, 0.170) | 0.236 | **0.148 (0.027, 0.246)** | **0.007** |
| | | $W_{LE}$ | 0.026 (-0.052, 0.130) | 0.599 | 0.030 (-0.043, 0.106) | 0.630 |
| | *Plastic & irreversible* | $D_1$ | 0.020 (-0.055, 0.100) | 0.723 | 0.064 (-0.007, 0.198) | 0.081 |
| | | $D_R$ | -0.010 (-0.076, 0.082) | 0.963 | 0.052 (-0.005, 0.161) | 0.084 |
| | | $W_D$ | 0.018 (-0.054, 0.078) | 0.885 | 0.016 (-0.040, 0.100) | 0.629 |
| | | $W_R$ | 0.045 (-0.057, 0.173) | 0.492 | 0.018 (-0.031, 0.162) | 0.437 |
| **Combined Descriptors** | *Strength-Adjusted for* | $F_U + D_U$ | 0.017 (-0.062, 0.129) | 0.620 | 0.106 (-0.004, 0.247) | 0.061 |
| | | $F_U + W_U$ | 0.032 (-0.057, 0.159) | 0.479 | **0.126 (0.006, 0.281)** | **0.035** |
| | | $F_U + D_1$ | 0.018 (-0.066, 0.119) | 0.684 | 0.097 (-0.007, 0.231) | 0.072 |
| | | $F_U + D_R$ | 0.005 (-0.066, 0.101) | 0.893 | 0.085 (-0.005, 0.220) | 0.070 |
| | | $F_U + W_D$ | 0.009 (-0.064, 0.114) | 0.726 | 0.120 (-0.011, 0.238) | 0.087 |
| | | $\boldsymbol{F_U + W_R}$ | 0.114 (-0.002, 0.229) | 0.053 | **0.202 (0.075, 0.333)** | **p < 0.001** |
| | *Linear Elastic Force-Adjusted for* | $\boldsymbol{F_{LE} + D_{LE}}$ | 0.090 (-0.023, 0.191) | 0.129 | **0.161 (0.046, 0.307)** | **0.002** |
| | | $\boldsymbol{F_{LE} + W_{LE}}$ | 0.093 (-0.004, 0.211) | 0.060 | **0.182 (0.071, 0.326)** | **0.001** |
| | | $F_{LE} + D_1$ | 0.029 (-0.061, 0.120) | 0.603 | 0.121 (-0.006, 0.234) | 0.071 |
| | | $F_{LE} + D_R$ | 0.002 (-0.064, 0.123) | 0.759 | 0.112 (-0.004, 0.230) | 0.065 |
| | | $F_{LE} + W_D$ | 0.054 (-0.049, 0.146) | 0.462 | 0.117 (-0.002, 0.258) | 0.055 |
| | | $\boldsymbol{F_{LE} + W_R}$ | 0.123 (-0.016, 0.263) | 0.099 | **0.187 (0.047, 0.355)** | **0.002** |

*NRI was computed by adding each parameter to the reference models (TH aBMD or FN aBMD), to assess their added predictive value. **P-values were derived from two-sided bootstrap resampling of the total NRI estimate; bold descriptor names indicate selection based on AUROC performance (Table 2), whereas bold NRI values denote statistically significant improvement in predictive performance relative to TH aBMD or FN aBMD.

To assess the added predictive value beyond DXA, integrated discrimination improvement (IDI) and continuous NRI (cNRI) were evaluated for 3D-DXA–based parameters and FE-based descriptors, using TH aBMD and FN aBMD as reference models (Table D3).

Table D3. IDI and cNRI for DXA, 3D-DXA–based parameters, and FE-based descriptors vs. TH and FN aBMD

| Methods | | | Descriptor | Vs TH aBMD as a reference | | Vs FN aBMD as a reference | | Vs TH aBMD as a reference | | Vs FN aBMD as a reference | |
|---|---|---|---|---|---|---|---|---|---|---|---|
| | | | | IDI* (95% CI) | *P*-Value** | IDI* (95% CI) | *P*-Value** | cNRI* (95% CI) | *P*-Value** | cNRI* (95% CI) | *P*-Value** |
| **DXA** | | | TH aBMD | Ref | - | - | | Ref | - | - | - |
| | | | FN aBMD | - | - | Ref | | - | - | Ref | - |
| **3D-DXA–Based Parameters (TH)** | | | Int.vBMD | **0.013 (0.001, 0.041)** | **0.01** | **0.016 (0.002, 0.048)** | **0.013** | **0.315 (0.063, 0.537)** | **0.015** | **0.298 (0.016, 0.563)** | **0.035** |
| | | | Trab.vBMD | **0.027 (0.006, 0.067)** | **0.003** | **0.029 (0.006, 0.069)** | **0.003** | **0.579 (0.322, 0.754)** | **p < 0.001** | **0.488 (0.195, 0.771)** | **p < 0.001** |
| | | | Cort.sBMD | 0.001 (-0.000, 0.015) | 0.261 | 0.002 (-0.000, 0.016) | 0.232 | -0.060 (-0.214, 0.272) | 0.937 | 0.145 (-0.078, 0.339) | 0.237 |
| **FE-Based Descriptors (TH)** | **Nonlinear FE Analysis** | | $F_U$ | -0.000 (-0.001, 0.006) | 0.507 | 0.001 (-0.001, 0.012) | 0.372 | -0.016 (-0.088, 0.303) | 0.354 | 0.130 (-0.088, 0.372) | 0.265 |
| | | | $D_U$ | 0.007 (-0.001, 0.030) | 0.188 | 0.012 (-0.001, 0.043) | 0.083 | **0.291 (0.049, 0.493)** | **0.025** | **0.402 (0.144,0.611)** | **p < 0.001** |
| | | | $W_U$ | 0.004 (-0.001, 0.020) | 0.181 | 0.002 (-0.001, 0.018) | 0.276 | 0.192 (-0.034, 0.430) | 0.108 | 0.197 (-0.043, 0.422) | 0.111 |
| | **Piecewise Linear Analysis** | *Elastic* | $F_{LE}$ | -0.000 (-0.001, 0.007) | 0.591 | 0.002 (-0.001, 0.014) | 0.317 | 0.064 (-0.105, 0.306) | 0.341 | 0.168 (-0.058, 0.385) | 0.167 |
| | | | $D_{LE}$ | **0.025 (0.001, 0.066)** | **0.033** | **0.030 (0.002, 0.077)** | **0.019** | **0.332 (0.091, 0.550)** | **0.005** | **0.368 (0.155, 0.667)** | **0.001** |
| | | | $W_{LE}$ | 0.009 (-0.000, 0.037) | 0.082 | 0.003 (-0.001, 0.022) | 0.267 | 0.162 (-0.080, 0.441) | 0.193 | 0.196 (-0.066, 0.418) | 0.172 |
| | | *Plastic & irreversible* | $D_1$ | 0.003 (-0.001, 0.018) | 0.320 | 0.007 (-0.001, 0.030) | 0.130 | 0.170 (-0.053, 0.414) | 0.135 | **0.344 (0.068, 0.571)** | **0.017** |
| | | | $D_R$ | 0.002 (-0.001,0.016) | 0.393 | 0.006 (-0.001, 0.027) | 0.184 | 0.118 (-0.093, 0.331) | 0.298 | 0.209 (-0.011, 0.437) | 0.068 |
| | | | $W_D$ | 0.002 (-0.001, 0.015) | 0.271 | 0.001 (-0.001, 0.014) | 0.340 | 0.059 (-0.100, 0.304) | 0.379 | 0.101 (-0.091, 0.347) | 0.256 |
| | | | $W_R$ | 0.011 (-0.000, 0.040) | 0.052 | 0.005 (-0.001, 0.028) | 0.185 | **0.238 (0.006, 0.555)** | **0.043** | 0.170 (-0.045, 0.400) | 0.161 |
| | **Combined Descriptors** | *Strength-Adjusted for* | $F_U + D_U$ | 0.006 (-0.001, 0.030) | 0.125 | **0.013 (0.001, 0.045)** | **0.031** | **0.370 (0.139, 0.661)** | **0.005** | **0.418 (0.122, 0.662)** | **0.005** |
| | | | $F_U + W_U$ | 0.007 (-0.001, 0.029) | 0.076 | **0.013 (0.001, 0.042)** | **0.024** | **0.456 (0.216, 0.757)** | **p < 0.001** | **0.494 (0.167,0.716)** | **p < 0.001** |
| | | | $F_U + D_1$ | 0.003 (-0.001, 0.020) | 0.215 | 0.008 (-0.000, 0.034) | 0.057 | **0.345 (0.032, 0.608)** | **0.03** | **0.319 (0.057, 0.604)** | **0.016** |
| | | | $F_U + D_R$ | 0.002 (-0.001, 0.017) | 0.238 | 0.007 (-0.001, 0.030) | 0.088 | 0.332 (-0.059, 0.556) | 0.125 | **0.274 (0.040, 0.508)** | **0.025** |

Table D3. (continued)

| Methods | Descriptor | Vs TH aBMD as a reference | | Vs FN aBMD as a reference | | Vs TH aBMD as a reference | | Vs FN aBMD as a reference | |
|---|---|---|---|---|---|---|---|---|---|
| | | IDI* (95% CI) | $P$-Value** | IDI* (95% CI) | $P$-Value** | cNRI* (95% CI) | $P$-Value** | cNRI* (95% CI) | $P$-Value** |
| | $F_U + W_D$ | 0.003 (-0.001, 0.020) | 0.177 | 0.008 (-0.000, 0.032) | 0.066 | **0.356 (0.009, 0.624)** | **0.046** | **0.291 (0.062, 0.527)** | **0.015** |
| | **$F_U + W_R$** | **0.036 (0.009, 0.079)** | **p < 0.001** | **0.042 (0.012, 0.090)** | **p < 0.001** | **0.570 (0.361, 0.795)** | **p < 0.001** | **0.459 (0.220, 0.761)** | **p < 0.001** |
| *Linear Elastic Force-Adjusted for* | **$F_{LE} + D_{LE}$** | **0.028 (0.003, 0.070)** | **0.013** | **0.033 (0.005, 0.083)** | **0.006** | **0.465 (0.230, 0.698)** | **p < 0.001** | **0.399 (0.160, 0.690)** | **0.001** |
| | **$F_{LE} + W_{LE}$** | **0.033 (0.008, 0.070)** | **p < 0.001** | **0.038 (0.010, 0.084)** | **p < 0.001** | **0.542 (0.354, 0.769)** | **p < 0.001** | **0.488 (0.241, 0.742)** | **p < 0.001** |
| | $F_{LE} + D_1$ | 0.003 (-0.001, 0.020) | 0.206 | **0.009 (0.000, 0.033)** | **0.049** | **0.380 (0.049, 0.603)** | **0.026** | **0.362 (0.072, 0.616)** | **0.015** |
| | $F_{LE} + D_R$ | 0.002 (-0.001, 0.018) | 0.256 | 0.007 (-0.000, 0.031) | 0.073 | 0.419 (-0.078, 0.586) | 0.109 | **0.245 (0.035, 0.536)** | **0.020** |
| | $F_{LE} + W_D$ | 0.004 (-0.001, 0.021) | 0.15 | **0.009 (0.000, 0.033)** | **0.035** | **0.431 (0.105, 0.671)** | **0.0130** | **0.384 (0.110, 0.640)** | **0.006** |
| | **$F_{LE} + W_R$** | **0.026 (0.006, 0.067)** | **0.002** | **0.031 (0.008, 0.075)** | **p < 0.001** | **0.476 (0.222, 0.734)** | **p < 0.001** | **0.435 (0.231, 0.750)** | **p < 0.001** |

*IDI and cNRI were computed by adding each parameter to the reference models (TH aBMD or FN aBMD), to assess their added predictive value. **P-values were derived from two-sided bootstrap resampling; bold descriptor names indicate selection based on AUROC performance (Table 2), whereas bold IDI and cNRI values denote statistically significant improvement in predictive performance relative to TH aBMD or FN aBMD.

Table D4 summarizes HRs with 95% CIs per one SD change for DXA, 3D-DXA–based parameters, and FE-based descriptors.

Table D4. HRs with 95% CI per one SD change for DXA, 3D-DXA–based parameters, and FE-based descriptors.

| | | | | HR (95% CI) | $P$-Value** |
|---|---|---|---|---|---|
| **DXA** | | | TH aBMD | 2.696 (2.289, 3.223) | <0.001 |
| | | | FN aBMD | 2.746 (2.299, 3.316) | <0.001 |
| **3D-DXA–Based Parameters (TH)** | | | Int.vBMD | **2.978 (2.485, 3.638)** | <0.001 |
| | | | Trab.vBMD | **3.268 (2.702, 4.035)** | <0.001 |
| | | | Cort.sBMD | 2.515 (2.121, 3.026) | <0.001 |
| **FE-Based Descriptors (TH)** | **Nonlinear FE Analysis** | | $F_U$ | 2.693 (2.228, 3.291) | <0.001 |
| | | | $D_U$ | 1.941 (1.656, 2.293)* | <0.001 |
| | | | $W_U$ | 1.839 (1.483, 2.296) | <0.001 |
| | **Piecewise Linear Analysis** | *Elastic* | $F_{LE}$ | **2.753 (2.281, 3.362)** | <0.001 |
| | | | $D_{LE}$ | 1.797 (1.605, 2.092)* | <0.001 |
| | | | $W_{LE}$ | 2.034 (1.632, 2.550) | <0.001 |
| | | *Plastic & irreversible* | $D_1$ | 1.834 (1.574, 2.149)* | <0.001 |
| | | | $D_R$ | 1.762 (1.513, 2.050)* | <0.001 |
| | | | $W_D$ | 1.829 (1.482, 2.277) | <0.001 |
| | | | $W_R$ | 1.874 (1.509, 2.360) | <0.001 |
| | **Combined Descriptors** | *Strength-Adjusted for* | $F_U + D_U$ | 2.236 (1.794, 2.827) | <0.001 |
| | | | $F_U + W_U$ | **3.229 (2.610, 3.959)** | <0.001 |
| | | | $F_U + D_1$ | 2.282 (1.830, 2.867) | <0.001 |
| | | | $F_U + D_R$ | 2.336 (1.885, 2.940) | <0.001 |
| | | | $F_U + W_D$ | **3.021 (2.445, 3.729)** | <0.001 |
| | | | $\mathbf{F_U + W_R}$ | **3.577 (2.821, 4.597)** | <0.001 |
| | | *Linear Elastic Force-Adjusted for* | $\mathbf{F_{LE} + D_{LE}}$ | 2.421 (1.987, 3.028) | <0.001 |
| | | | $\mathbf{F_{LE} + W_{LE}}$ | **3.330 (2.623, 4.281)** | <0.001 |
| | | | $F_{LE} + D_1$ | 2.334 (1.876, 2.941) | <0.001 |
| | | | $F_{LE} + D_R$ | 2.397 (1.942, 3.016) | <0.001 |
| | | | $F_{LE} + W_D$ | **3.109 (2.501, 3.858)** | <0.001 |
| | | | $\mathbf{F_{LE} + W_R}$ | **3.541 (2.797, 4.515)** | <0.001 |

* HR for 1 SD increase, for other values without *, the HR results are for 1 SD decrease. ** P-Values were obtained via nonparametric bootstrap resampling; Bold descriptors reflect AUROC-based selection (Table 2), and bold HR values indicate estimates greater than TH aBMD.
